# Target Trial Emulation Applications in Hypertension Research: A Scoping Review

**DOI:** 10.1101/2025.04.21.25326150

**Authors:** Amir Habibdoust, Hanxiao Zuo, Richelle J. Koopman, Aditi Gupta, Diego R. Mazzotti, Xing Song

**Affiliations:** Institute for Data Science and Informatics, University of Missouri-Columbia, Missouri, USA; School of Public Health, University of Alberta, Edmonton, Alberta, Canada; Department of Family and Community Medicine, University of Missouri-Columbia, Missouri, USA; Department of Internal Medicine, University of Kansas Medical Center, Kansas City, KS, USA; Division of Nephrology and Hypertension, Department of Internal Medicine, University of Kansas Medical Center, Kansas City, KS, USA; Division of Medical Informatics, Department of Internal Medicine, University of Kansas Medical Center, Kansas City, KS, USA; Division of Pulmonary Critical Care and Sleep Medicine, Department of Internal Medicine, University of Kansas Medical Center, Kansas City, KS, USA; Department of Biomedical Informatics, Biostatistics, and Medical Epidemiology, University of Missouri-Columbia, Missouri, USA

**Keywords:** Target Trial Emulation, Hypertension, Observational Data, Heterogeneous Treatment Effect, Scoping Review

## Abstract

**Objectives:** Target Trial Emulation (TTE) has emerged as a rigorous framework for causal inference using observational data, but its application in hypertension research remains underexplored. This scoping review aims to map current TTE applications, identify methodological strengths and weaknesses, and propose future directions for its use in hypertension research.

**Study Design and Setting:** We performed a scoping review following the Joanna Briggs Institute (JBI) guidance and the PRISMA extension for Scoping Reviews (PRISMA-ScR) checklist. We searched multiple databases, and three independent reviewers conducted screening and extraction using Covidence review management software. 14 out of 1,352 articles met the inclusion criteria.

**Results:** Most studies used data from electronic health records, claims databases, and registries. All of the interventions were pharmacological except for one. Common confounding adjustment methods included inverse probability weighting (50%) and the g-formula (21.5%), complemented by regression-based models. However, time-varying confounders were inconsistently addressed, and loss to follow-up was often managed through simple censoring rather than statistical methods. Residual confounding remained a concern—although several studies acknowledged unobserved confounders, only five (36%) employed negative controls or e-values to assess their impact. While subgroup analyses were common, explicit heterogeneous treatment effect (HTE) estimation was limited. Advanced causal machine learning techniques for bias mitigation or HTE detection were not reported.

**Conclusion:** TTE shows strong potential to complement randomized controlled trials in hypertension research by providing more generalizable insights. While still in its early stages, current studies highlight its ability to address key challenges such as HTE, long-term outcomes, and dynamic treatment strategies.

## 1. Introduction

Hypertension is a considerable risk factor associated with a range of health issues, including cardiovascular disease, stroke, chronic kidney disease, and cognitive decline^1–3^ and is a leading preventable risk factor for global mortality^4^. Despite advancements in the availability of effective and safe antihypertensive medications^5^, globally only 23% of treated women and 18% of treated men achieve targeted pressure levels^6^. While there has been significant progress in our understanding of blood pressure, challenges and open questions still persist in the field^7–9^, such as, causes of hypertension^10^, the heterogeneous effectiveness and safety of different drug classes across diverse groups (ranging from children to older adults, and across different races, sexes, and comorbidities)^8,11,12^, blood pressure variability^13^, and barriers and promoters to short-term and long-term adherence to hypertension treatment^8^.

However, many questions are not infeasible or impractical to be addressed with randomized controlled trials (RCTs), due to high time and effort consumption, high costs, high compliance burden for participants, low tolerance of deviation from the study protocol^12^, to name a few. The constricted patient selection criteria of many RCTs and other limiting recruitment factors (e.g., geography) can lead to a significant lack of generalizability (external validity)^14–16^, and hinder the assessment of complex and heterogeneous treatment effects^8,11,12,17–20^. While pragmatic trials may enhance real-world applicability to some extent, interventional RCTs remain constrained by several key limitations^21,22^. These include ethical concerns^23,24^ (e.g., the use of placebos and certain treatments that may be less desirable for older adults and ^25–29)^, as well as challenges in evaluating dynamic, time-varying treatment effects^9,30,31^, lifelong management^8^; and long-term treatment effects^8^ of hypertension interventions.

Hence, large-scale, real-world, observational data provides a promising, alternative avenue for estimating causal treatment effects in more generalizable settings, at much lower costs, for longer term^23,24^. The rapid growth in the use of the internet, social media, digital health technologies such as biosensors, mobile and wearable devices, as well as claims and billing activities, disease registries, and electronic health records (EHRs), together with increased capacity in data storage and computation, have led to the accumulation of large amounts of digital real-world data. However, as a result of lack of rigor in the non-research-specific data collection process, estimating CTE from observational real-world data (RWD) is not straightforward and often leads to various sources of bias, such as, selection bias, immortal time bias, measurement bias, confounding bias (omitted confounders, unobserved confounders, time-varying confounding), and reverse causality.^24,32–35^

In response to these challenges, Target Trial Emulation (TTE) has emerged as a promising framework for CTE estimation using observational data. Following the “potential outcome” framework^36,37^, TTE attempts to mimic RCTs by sampling the observational data in a deliberate fashion that closely follows RCT protocols. Although it cannot fully eliminate all biases (such as unmeasured confounders), TTE offers systematic approach to reduces biases related to erroneous study designs including selection and immortal time bias^38–40^, making it especially valuable in hypertension research. Despite the rapid increasing of TTE in medical literature in recent years, its adoption into hypertension research remains limited compared to fields like cancer or other cardiovascular research^41^. For those that claim to use TTE may not necessarily apply the framework correctly^41^.

Our scoping review aimed to scan existing literature on applications of the TTE in hypertension research to answer the primary question: “how has TTE been applied in hypertension research?”, which we further break down into: a) what types of questions have been addressed using TTE in hypertension research (e.g., comparative effectiveness of different treatments); b) what types of study designs have been effectively emulated; c) what types of biases were identified using TTE and how they were addressed (e.g., selection bias, censoring bias, confounding bias); d) whether and how studies examined heterogeneous treatment effects (HTE); e) how well the study follow the TTE framework and where the deviations occurred; f) what types of statistical or machine learning methods were applied for causal effect estimation. Based on our findings, we will offer methodological recommendations and discuss how TTE can address critical gaps in hypertension research. In addition, we hope that this review will encourage researchers to appropriately adopt the TTE framework and raise awareness of new opportunities and relevant datasets for hypertension research.

## 2. Methods

The protocol of this scoping review was registered in Open Science Framework ^42^. We followed the Joanna Briggs Institute (JBI) guidance^43^ to conduct our scoping review and used Preferred Reporting Items for Systematic reviews and Meta-Analyses extension for Scoping Reviews (PRISMA-ScR) checklist to ensure the rigor of this study and methodological transparency ^44^ (see Supplement Appendix A for the full protocol).

### 2.1. Eligibility Criteria, Search strategy, and Screening

We searched PubMed, Google Scholar, and Web of Science databases from January 2015 to November 18, 2024. We used multiple synonymic words or phrases representing the concept “target trial emulation" and "hypertension" as key search terms (see Supplement Appendix B). All retrieved papers were exported to COVIDENCE review management software^45^. Duplicates removal and screening were aided by COVIDENCE. After removing duplicates, three independent reviewers manually reviewed (A.H., H.Z., and X.S.), the titles and abstracts based on a consensus set of inclusion criteria: a) article type should be original research published in a scientific peer-reviewed journals; b) study designs should follow TTE framework at least to some extent if not entirely; c) written in English; d) the primary objective of the study should be related to hypertension and blood pressure control. Disagreements were addressed and resolved by having a discussion and majority voting during the review process. Finally, all remaining articles satisfying the inclusion criteria were fully reviewed and analyzed.

### 2.2. Data extraction

We extracted the following information from each study: (1) Eligibility criteria and data source, (2) Treatment strategies and assignment, (3) Outcomes, (4) Bias sources, (5) Causal contrast strategy, (6) Analysis plan and statistical methods, and (7) Limitations. The data were independently extracted by the three reviewers (A.H., H. Z., and X.S.) and discrepancies resolved through consensus. The main idea of this extraction categorization is based on the target trial emulation design components proposed by Hernán and Robins (2016)^24^, which include eligibility criteria, treatment strategy, assignment procedure, follow-up, primary outcome definition, the causal contrast of interest, and a statistical analysis plan. Each study’s design was assessed against these components. Additionally, we incorporated specific questions and categories to focus on different sources of bias. Supplemental Appendix provides a detailed overview of the questions used to evaluate each study.

## 3. Results

### 3.1. Study characteristics

A total of 1352 articles were retrieved from the three selected databases, with 744 duplicates removed. After screening, 14 articles met the inclusion criteria^1^ and were included in this review (see Figure 1). Of the selected studies, most were conducted in the USA (n = 9), with others from Sweden (n = 2), the Netherlands (n = 1), and the UK (n = 2), all published between 2021 and 2024.

**Figure 1.**
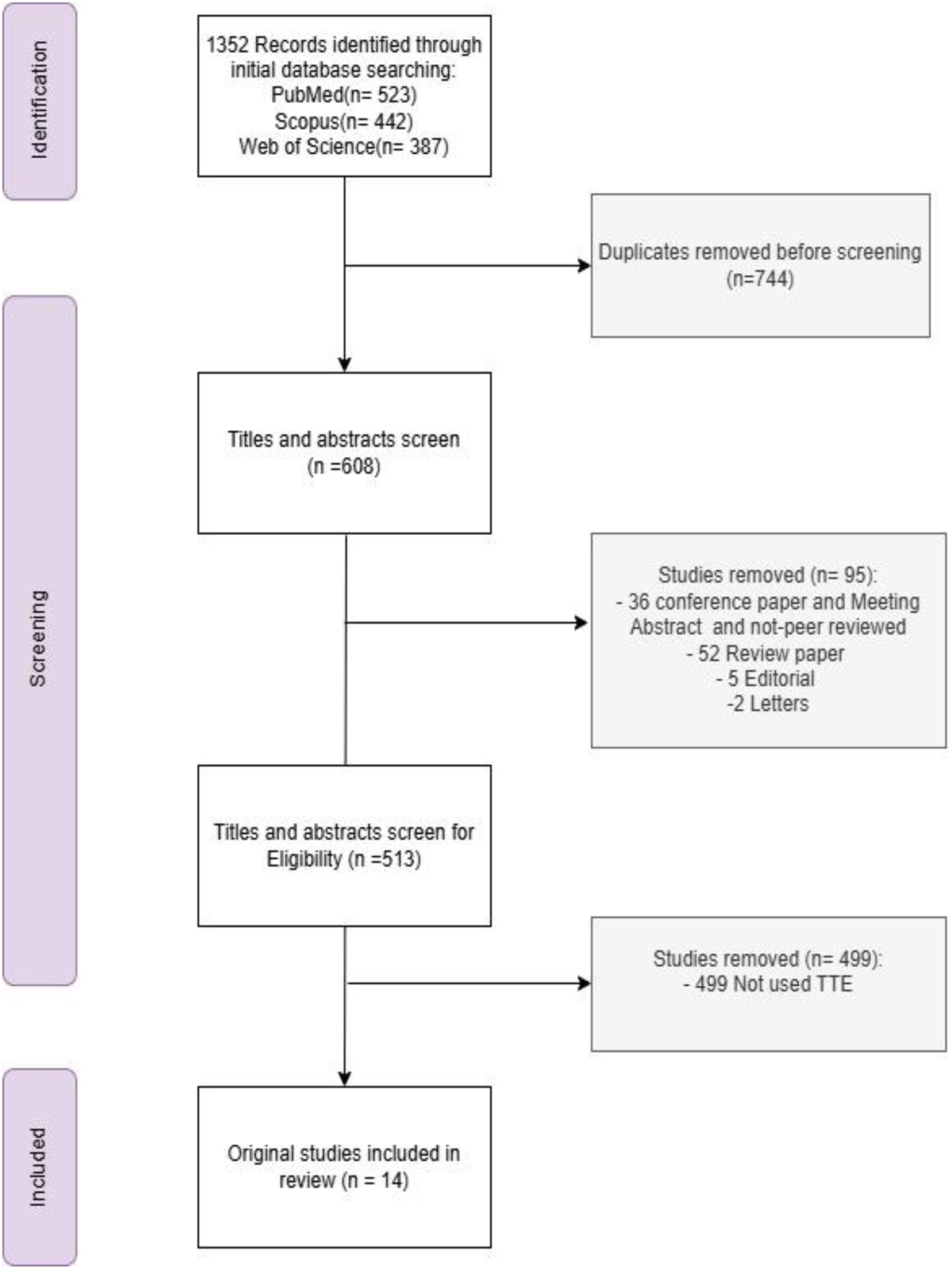
PRISMA Flowchart.

The data sources in the USA were Veterans Health Administration (VHA), Medicare, Health and Retirement Study (HRS), Multi-Ethnic Study of Atherosclerosis (MESA), Electronic Health Record (EHR), and the IBM LCED database. In the UK, the data source was Clinical Practice Research Datalink (CPRD) and CPRD Aurum (EHR), in the Sweden combination of Electronic Health Record System and National Patient Register, Swedish Mammography Cohort (SMC) and the Cohort of Swedish Men (COSM). In the Netherlands, the data was linked between the Rotterdam Study (a prospective cohort study) and Regional Institute for Outpatient Mental Health Care dataset.

The sample size and target population varied across the studies. Seven studies included 1,000 to 100,000 subjects^47–53^, five studies had between 100,000 and 200,000 subjects^31,54–57^, and two studies had a sample size of more than 200,000^54,58^. Regarding the target population, five studies were conducted among veterans, three focused on adults in midlife, three on those over 65, one on those older than 50, two on individuals over 18, and one on those between 55 and 88 years old.

### 3.2. Research Focus, Treatment, and Outcome

Guided by our research question, we focused on the domain of hypertension. Accordingly, the we categorized our research questions into two broad categories: 1) Comparative effectiveness of different hypertension treatment strategies and management approaches (7 studies) ^31,46,52,54,55,57^; and 2) short- or long-term impact of high or uncontrolled BP (7 studies) (^47–50,56,58,59^). We also included a study that employed the TTE framework to explore whether existing antihypertensive medications can be repurposed to treat other health conditions^53^. Table 1 summarizes the important findings. We also included a study that employed the TTE framework to explore whether existing antihypertensive medications can be repurposed to treat other health conditions (^53^).

**Table 1.**
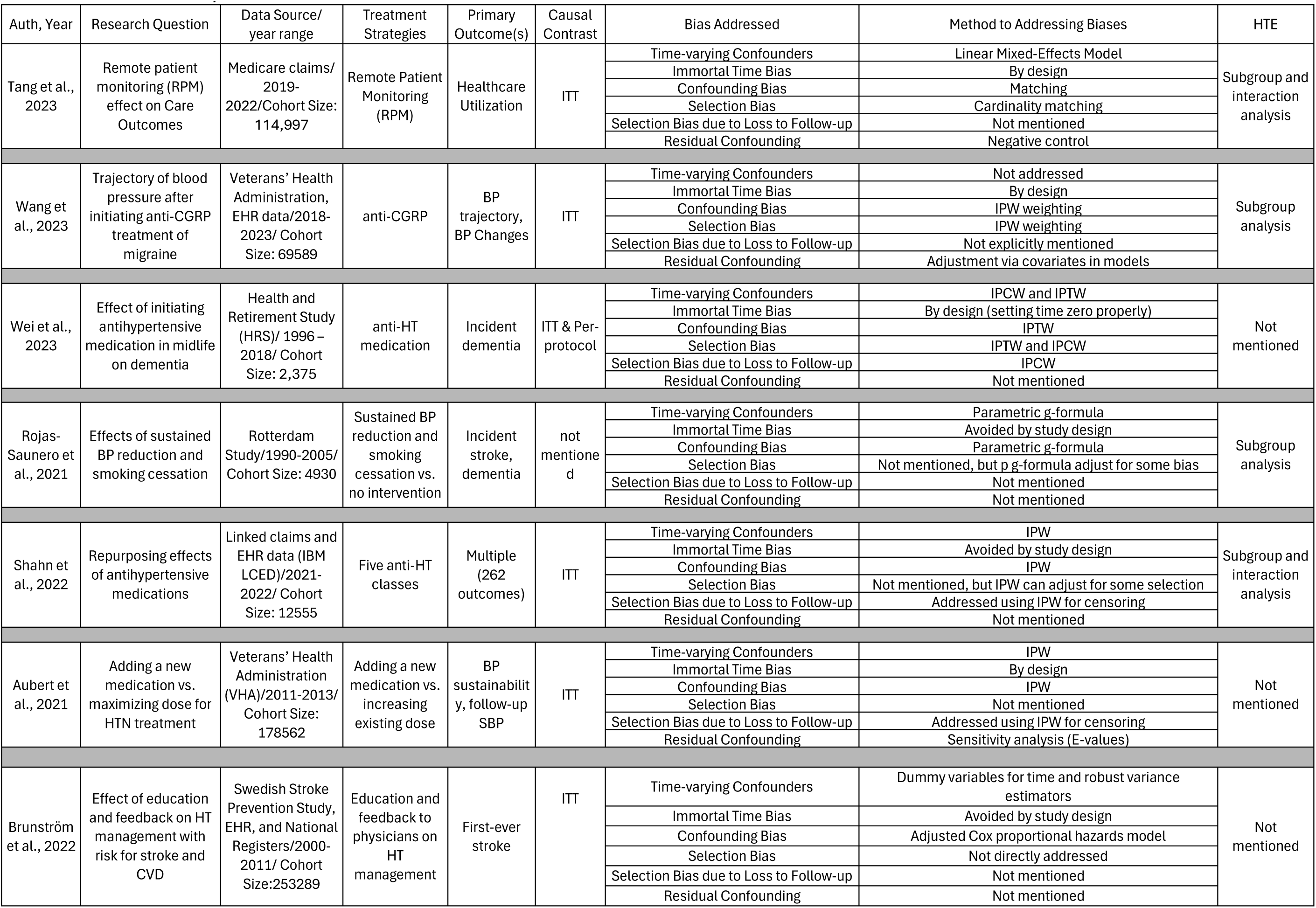
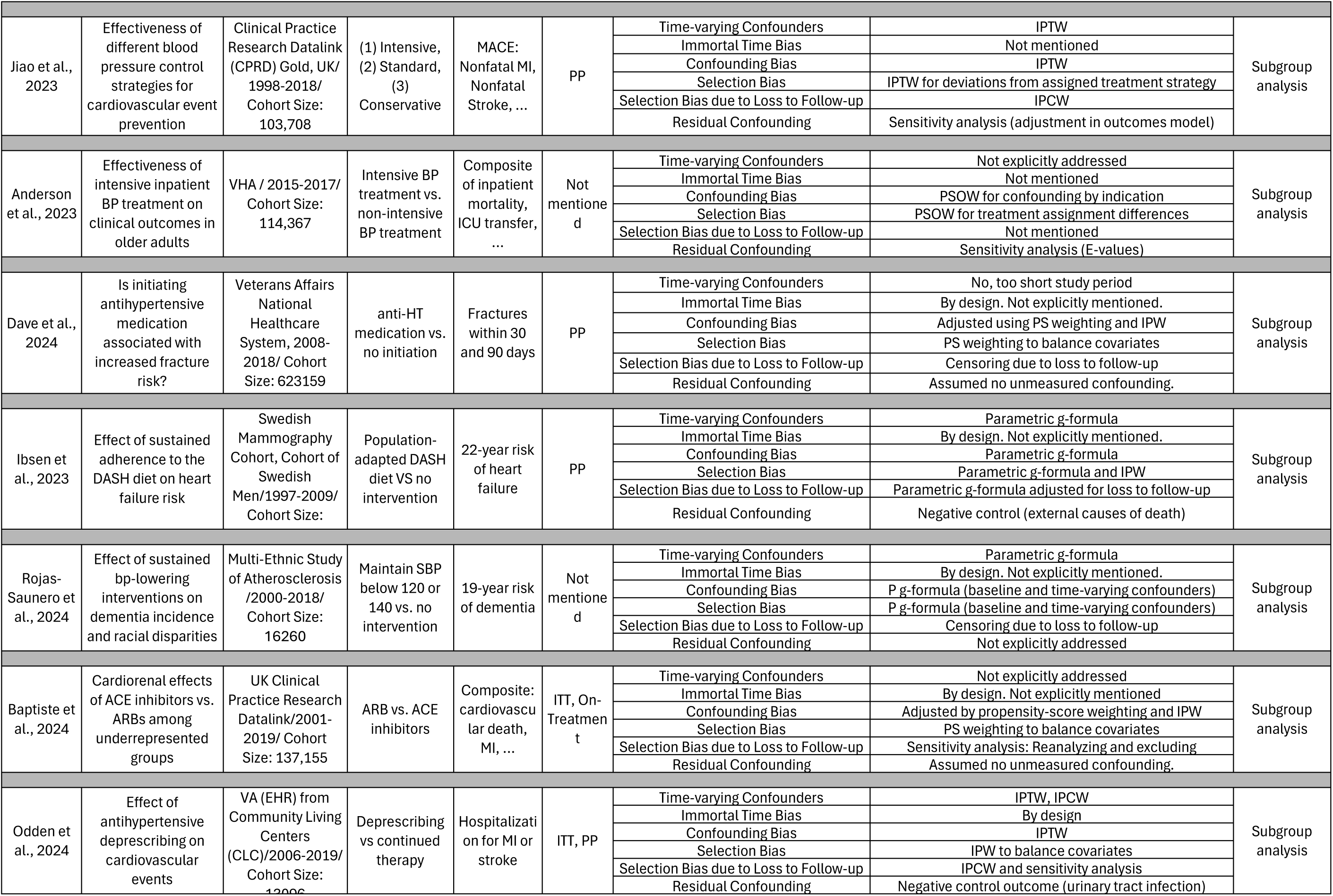
Summary of Extracted Information.

A variety of treatment strategies and management approaches have been examined, including: remote patient monitoring(^57^) and educational intervention(^54^) (2 studies), intervention time point(^48^) (1 study), sustained^49,59^ and adaptive^31^ blood pressure control (3 studies), diet intervention (^50^) (1 study), different class of antihypertensive drugs(^56,58^) (2 study), new medication versus maximizing dose(^46^) (1 study), deprescribed or continue antihypertensive therapy^52^ (1 study)and intensive antihypertensive treatment(^55^) (1 study) (See Table 1). One TTE study^56^ was conducted to replicate the reference RCT while also investigating the generalizability of the treatments of interests.

In terms of outcome, studies are focused on change in systolic blood pressure (SBP) (^46,47^) (2 studies), dementia and cognitive decline outcomes (^48,49,51^) (3 studies), healthcare utilization outcomes (^57^). Other Diseases Risk and other events Reduction (Cardiovascular, acute kidney injury (AKI), stroke, hospitalization, spending, falls, and etc.) Outcomes (^31,49,50,52,54–56,58^)(8 studies). Studies had different measurement approaches for outcome assessment inducing continuous, ordinal, binary, and time-to-event.

### 3.3. Reference RCT Type

It is noteworthy that four studies emulated a single arm RCT (^48,48,54,58^), while the remaining studies mimicked multiple-arm RCTs. In addition, one study emulated clustered RCT (^57^), where groups (or clusters) rather than individuals are randomized; another mimicked an adaptive RCT (^31^), a type that allows modifications to the trial procedures, and the remaining studies emulated a standard RCT.

### 3.4. Treatment Effect Estimation

Regarding causal contrast of interest, seven studies employed intention to treat (ITT)(^46,47,51,53,54,56,57^), four studies used per protocol (PP)(^31,49,50,58^), and two studies utilized both methods (^48^), and one study did not explicitly mention using ITT, but based on the study design, it appears they likely did. (^55^)(Table 1). Mean difference, as effect size measure, was used to quantify the causal contrast of interest in three studies (^46,47,57^), Risk ratio(RR) was used in three studies(^30,31,36^), three studies used hazard ratio(HR) as effect size(^31,52,54,56,58^), one study employed odds ratio(^55^), and one study reported both HR and RR(^53^), one employed both risk difference and risk ratio(^50^), one used both percentage and mean differences(^46^).

### 3.5. Design-Based Biases

#### 3.5.1. Sample and Immortal Time Biases

All final selected studies followed the TTE framework except one study did not include full implementation details on all the emulation and analytical steps (^46^). All studies tried to address immortal time bias by carefully defining baseline and inclusion criteria.

#### 3.5.2. Follow-up and related Biases

The studies covered a wide range of follow-up periods, depending on the research question, from less than 30 days (^58^) to 22 years (^50^). Decisions about the approach to loss to follow-up is a common challenge, particularly when blood pressure is the outcome, due to the lack of measurements for a considerable number of patients. Studies that followed a per-protocol (PP) analysis naturally drop or censor patients who did not have blood pressure measurements at the follow-up date. However, one study employed the g-formula to handle missing data and avoid bias, rather than simply excluding those cases (^50^).

Attrition bias (i.e., bias introduced by loss of follow-up) in TTE can be approached using several methods^60–63^. One study employed stabilized inverse probability of treatment weighting (IPTW) to address biases related to loss to follow-up (^53^); three study employed time-varying stabilized inverse probability of censoring weights (IPCW) (^31,48,52^); one studies used IPW (^46^); and one study applied parametric g-formula adjusted for loss to follow-up (^50^). Eight studies did not address attrition bias (simply censored) or explicitly did not mention it (^47,49,51,54,55,57,58^).

However, the study (^57^) that emulated a clustered RCT accounted for some loss to follow-up through censoring, implicitly assuming that censoring is non-informative^61^, which may reduce the relevance of bias associated with this issue. Ideally, though, bias from loss to follow-up should be addressed directly^61^ (See Table 1).

Specifically, regarding the loss to follow up due to lack of BP measurement before the end of the follow-up period, one study excluded patients without follow-up blood pressure measurements were from the analysis (^47^), by emphasizing independence from the outcome and using complete case analysis. Another study calculated the mean within 3 to 12 months, and if they could not find any data during these months, they addressed this loss by IPCW (^46^).

#### 3.5.3. Confounding Biases

In addressing confounding and emulating certain aspects of random assignment, various statistical methods were employed across the studies. Seven studies used IPTW (^31,46–48,52,53,56^), while three studies employed the g-formula method (^49–51)^. Two studies utilized regression techniques (^48,57^), one paper used propensity score overlap weighting (^55^), and one paper applied propensity score matching (^58^).

To address time-varying confounders, two studies employed regression methods (^56,57^), three studies used g-formula (^49–51)^, two studies applied stabilized IPTW (^31,52^). Four studies did not account for time-varying treatment and confounders (^47,48,53,55^), one study also did not consider the time varying issue due to short horizon (30 days) over which study outcomes were assessed (^58^). One study used a serial cohort design, including participants multiple times over the study period, which helped account for any time trends that might influence results (^54^) (See Table 1).

Regarding statistical methods to analyze outcomes, studies used one or a combination of different methods. Four studies used Cox regression model (^31,53,54,56,58^) and four studies applied logistic regression (^31,46,53,57^). Other methods included pooled logistics regression (^49,52^), weighted linear mixed-effects models (^47^), ordinary least squares regression(^57^), and overlap-weighted logistic regression(^55^) (See Table 1).

### 3.6. Heterogeneous treatment effect

In the medical field, heterogeneity of treatment effects (HTE) could be defined as the non-random variation in the direction or size of a treatment’s effect, across levels of a covariate against a clinical outcome, as measured by clinical outcomes^64,65^. To address HTE in clinical trials, traditional subgroup analyses are often theory-driven^66^, either pre-planned or post hoc. These approaches typically involve stratifying participants by pre-defined variables (e.g., age, sex, race) and testing for statistical significance of the interaction between the treatment and these stratifying variables^20,67,68^. While informative in some extent, such conventional approaches face several limitations. Each patient possesses multiple characteristics, resulting in a vast number of potential subgroups^69^. This introduces challenges such as increased risk of type I error due to multiple comparisons and reduced statistical power (type II error) for detecting true HTE^67,69^. Recently, more sophisticated, data-driven approaches have been developed to identify empirical HTE strata by leveraging machine learning models^20,66,68,70–74^.

Seven studies employed conventional subgroup (pre-specified) analysis as part of their sensitivity analysis (^49–52,55–57)^. One study found the results consistent across the pre-defined subgroups or little heterogeneity for primary outcome (^52,55,56^). In contrast, three studies found that heterogeneity across the pre-defined subgroups (^50,51,58^). The remaining six studies did not investigate the HTE (^31,46–48,53,54^) (See Table 1).

### 3.7. Study Acknowledged Limitations

We report only the limitations explicitly noted by the original authors, some of which were already addressed in the bias section above (Section 3.5). Certain limitations specific to the research questions of individual studies are not discussed here.

Unmeasured confounders and residual confounding were mentioned as limitations in eleven articles (^31,46–50,52–54,56,58^). Quality of data and measurement error were mentioned nine times as one of the limitations(^31,46–49,51,52,56,57^), limited generalizability was noted eight times(^46,51,52,54–58^), lack of blood pressure data(^47,49,52,56,57^) was identified five times, time varying treatment and confounders appeared in twice(^47,53^), detecting heterogeneity(^47,48,51,58^) appeared four times. Although unobserved confounders is always a concern in observational studies, three studies tried to directly address this issue (^46,52,55^).

## 4. Discussion

### Limited context of use

Our review indicates that TTE approaches have garnered growing interest in hypertension research, particularly since 2021, yet their application in this area remains less prevalent and diverse compared to other fields^41,75^. First, despite the well-recognized strength of real-world observational data, there are still very few studies that attempted to comprehensively evaluate the long-term neurological, cardiac, and renal impacts of uncontrolled blood pressure, or the long-term effectiveness and safety of antihypertensive drugs and other management strategies (e.g. remote monitoring, smoking cessation). Second, as a result of adherence challenges and time-varying nature of the hypertension, dynamic treatment regimens (DTRs)^76,77^ is another potential use context to evaluate adaptive management approaches (including timing of treatments). RCT designs (such as sequential multiple assignment randomized trials (SMARTs) and different adaptive designs, that have been developed to address DTRs are resource-intensive, require complex design and implementation, and carry a potential for dropouts and non-compliance. However, we found only one paper addressing an adaptive strategy^31^. Future work is needed to further broaden the applications of TTE approaches in hypertension research by investigating various adaptive management strategies under real-world settings that can better inform clinical practice for improving long-term outcomes.

### Limited focus on heterogeneous treatment effects

HTE is particularly relevant in blood pressure research as there is substantial variability in patients’ responses to medications (or other interventions) as well as in the short-term or long-term effects of uncontrolled blood pressure. While most RCTs can only focus on average treatment effects, TTE provides a unique opportunity to extensively evaluate HTE. Most of the selected TTE performed HTE analyses on a few pre-defined subgroups (e.g., based on age, sex, baseline BP, medication use). However, there remain opportunities to discover true high-risk and high-benefit subgroups through comprehensively leveraging all possible information (often high-dimensional data) available in the RWD sources. Future work is needed to explore more advanced methods (such as causal random forest) that will generate HTE-driven subgroups within the TTE framework to enhance our understanding and management of hypertension and the impact of blood pressure across diverse subgroups.

### Limited focus on guideline-specific emulations

In 2017, American College of Cardiology (ACC) and American Heart Association (AHA)^7^ developed new guidelines for prevention, detection, and management of high blood pressure in adults, highlighting the following major shifts: 1) more proactive and intensive management approach (e.g., targeting BP goals at 130/80, initiating antihypertensives; restarting antihypertensives for secondary stroke prevention); 2) treatment individualization; 3) recognition of nonpharmacological strategies; 4) advocating for remote monitoring and multidisciplinary care. ACC/AHA together with European Society of Hypertension (ESH)^78^ also identified the following research gaps: optimal and safe BP thresholds and targets in old and frail patients, optimal SBP and DBP level at different time points in life, choice of first-line antihypertensive agents, ambulatory or home BP measures across ethnicities, benefit of lifetime low BP, treatment of hypertension during pregnancy and related risks. Despite the significance of these issues, we found no studies that specifically addressed these gaps. Future work is needed to leverage TTE to provide timely evidence on the effectiveness of the guideline changes and address these well-recognized research gaps.

### Lack of replication studies

There is a lack of replication studies to validate the effectiveness of TTE by directly comparing within the hypertensive research domain. Only 1 reviewed study attempted to replicate the ON-TARGET trial but included limited discussion on comparisons between the trial results and TTE results^56^. One such effort is the RCT-DUPLICATE initiative (Randomized, Controlled Trials Duplicated Using Prospective Longitudinal Insurance Claims: Applying Techniques of Epidemiology) co-launched by FDA and NIH in 2017, which, however, did not strictly follow the TTE framework^79^. Future work is needed to conduct robust replication studies that inform the reliability and limitation of TTE methodologies.

### Lack of comprehensive approach on quantifying and mitigating biases

One of the biggest challenges in applying the TTE framework is the control of biases and estimation of their impact. All selected studies recognized multiple sources of bias (e.g., attrition bias, time-varying confounders, unmeasured confounding bias, measurement error, etc.), but only a subset of the studies primarily examined the unmeasured confounding bias via negative controls or E-value. Although several studies addressed immortal time bias ’by design’, few used causal diagrams to validate this. Such diagrams clarify whether bias stems from misclassification or selection— especially important in trials with time-varying exposure.^80^ Future work is needed to develop a robust framework to consistently identify and quantify biases that can be introduced at each TTE step, which can help with proper interpretation and use of the TTE results.

### Potential of integrating causal machine learning methods

All the reviewed articles used parametric statistical models for treatment effect estimation. While development of causal machine learning methods (e.g., causal forests, counterfactual modeling) has enhanced the causal inference and predictive inference^81,82^, it remain underutilized in detecting treatment heterogeneity in BP control contexts. None of the studies used any artificial intelligence (AI) and machine learning (ML) method in any steps of their study. Future work is needed to explore the wider adoption of advanced ML techniques to improve causal contrasting and counterfactual modeling.

### Limitations

The current scoping literature review is subject to a few limitations. First, we used several related keywords; however, since the TTE framework is rather new and still under development, researchers may use different terms to refer to TTE, leading to potential misses of relevant studies. Second, we are fully aware of the potential for publication bias, where studies with positive results are more likely to be published and thus selected in this review. Third, the subjective nature of study selection and data extraction between two reviewers may still introduce bias, particularly when disagreements arise that cannot be resolved, necessitating a judgment call; to help mitigate this bias, we used three reviewers.

## 5. Conclusion

This review emphasizes that TTE presents a promising framework for advancing hypertension research by utilizing large-scale, real-world observational data, particularly in situations where traditional RCTs encounter limitations and limited generalizability. While the applications of TTE in hypertension research is still in its early stage, current studies demonstrate its significant potential to complement RCT studies in addressing key challenges such as heterogenous treatment effect, long-term outcome, and dynamic treatment strategies. The paucity of available studies revealed in this scoping review emphasizes the need for more TTE investigations in this area, with the potential to help clinicians and patients improve hypertension management and outcomes in a timely fashion.

## Data Availability

All data produced in the present study are available online in the cited papers online.

## Footnotes

1 To ensure we did not overlook any studies that employed target trial emulation but did not mention it in the title or main text, we randomly sampled 20% of the papers and reviewed their full text. We did not find any such studies unless one^46^.

