## Supplementary material for "Target Trial Emulation Applications in Hypertension Research: A Scoping Review"

### Appendix

**Appendix A - Target Trial Emulation Application in the Area of Blood Pressure Research: a scoping review protocol**

**PROTOCOL DESIGN**

We followed the JBI guidance to design the protocol and conduct our scoping review of our scoping review[1] We also used Preferred Reporting Items for Systematic reviews and Meta-Analyses extension for Scoping Reviews (PRISMA-ScR) checklist to ensure the rigor of this study and methodological transparency[2].

**1- Identifying the research questions**

The PCC framework (population, concept, and context) was recommended as a guide for developing clear and meaningful objectives and eligibility criteria in a scoping review[3]. Hence, Using the Population, Concept, Context notation from the Joanna Briggs Institute (JBI)[1], we formulated our research question as below:

- **Concept**: Application of TTE for studying the hypertension treatment and effects of blood pressure.
- **Context**: Observational blood pressure research settings.

Since we have no limitation related to specific subgroups, our question is as follows:

- How has Target Trial Emulation (TTE) been applied in observational blood pressure research to study the effects of blood pressure on other diseases and the effectiveness of hypertension treatments?

**2- Inclusion Criteria**

Following Population, Concept, Context (PCC), our inclusion criteria are:

- Original research
- Published in Scientific Peer-reviewed
- Written in English
- Study designs based on target trial emulation framework
- Primary objects that include hypertension and blood pressure and impact of blood pressure

**3- Search strategy**

We searched PubMed, Google Scholar, and Web of Science from January 2015 to November 18, 2024. We used different keywords representing the concept “target trial emulation" and "blood pressure"(see Appendix B). All retrieved papers (1352) were exported to COVIDENCE[4] to review management software. Duplicates removal and screening were aided by COVIDENCE.

**4- Screening**

After removing duplicates, we reviewed the titles and abstract if the articles meet the inclusion criteria. The disagreements during the screening process of titles and abstracts were addressed and resolved by having a discussion between team members. Subsequently, all remaining articles that employed TTE in any blood pressure related issues, including blood pressure impact on health or hypertension treatment were independently and comprehensively reviewed by two reviewers.

##### 5- Data Extraction

We gathered the following details from each study: (1) Eligibility criteria and data sources, (2) Treatment approaches and assignment processes, (3) Outcomes, (4) Potential sources of bias, (5) Causal contrast approach, (6) Analysis plan and statistical methods, and (7) Study limitations. Three reviewers independently extracted the data, resolving any differences by consensus. This extraction framework is based on the target trial emulation components outlined by Hernán and Robins (2016)[5], Each study's design was evaluated against these components. In addition, we added specific questions and categories to address different bias sources and heterogeneous treatment effects. A detailed summary of the questions used for study evaluation can be found in Appendix C of this appendix.

##### 6- Analysis and Report

We reviewed the papers to understand how they apply the target trial emulation (TTE) framework in the context of blood pressure and hypertension. Specifically, our focus was on understanding how TTE was used in blood pressure research, including examining the effects of blood pressure on other diseases and the effectiveness of hypertension treatments. We also summarized how each study addressed various sources of bias, how AI or machine learning was used to enhance results, and any limitations identified. Finally, we discussed our findings, note research gaps, and provide suggestions for future research.

**Appendix B. Search terms and results of the literature review.**

| Search Statement | Medium | Time period | Retrieved papers |
|---|---|---|---|
| TITLE-ABS ("blood pressure" OR "Blood-Pressure" OR "Blood Pressure" OR "hypertension") AND TITLE-ABS ("real world data" OR "target trial emulation" OR "emulate target trial" OR "emulated target" OR "hypothetical trial" OR "hypothetical intervention" OR "hypothetical interventions") AND PUBYEAR > 2014 AND PUBYEAR < 2025 AND (LIMIT-TO (LANGUAGE, "English")) | Scopus | 01/01/2015 to 11/18/2025 | 442 |
| Title:<br>("blood pressure" OR hypertension OR "Blood-Pressure" OR "Blood-Pressure") AND (("real-world data") OR ("target trial emulation") OR ("emulate target trial") OR ("emulated target") OR ("hypothetical trial") OR ("hypothetical intervention") OR ("hypothetical interventions"))<br>OR Abstract:<br>("blood pressure" OR hypertension OR "Blood-Pressure" OR "Blood-Pressure") AND (("real-world data") OR ("target trial emulation") OR ("emulate target trial") OR ("emulated target") OR ("hypothetical trial") OR ("hypothetical intervention") OR ("hypothetical interventions")) | Web of Science | 01/01/2015 to 11/18/2025 | 387 |
| ((blood pressure[Title/Abstract]) OR (Blood-Pressure[Title/Abstract]) OR (Blood-Pressure[Title/Abstract]) OR (hypertension[Title/Abstract])) AND (("real-world data"[Title/Abstract]) OR (target trial emulation[Title/Abstract]) OR ("hypothetical trial"[Title/Abstract]) OR ("emulated target" [Title/Abstract]) OR ("hypothetical trial"[Title/Abstract]) OR ("hypothetical intervention"[Title/Abstract]) OR ("hypothetical interventions"[Title/Abstract]) OR(emulate target trial[Title/Abstract])) | PubMed | 01/01/2015 to 11/18/2025 | 523 |

* Results as of November 18, 2024

**Appendix C. Information extraction chart.**

| Questions | Response |
|---|---|
| What is the main question of the paper? | Text field |
| Publication year | Text field |
| Implementation country | Text field |
| **Eligibility Criteria and Data** | |
| What is the type of data source? | Real-world data (EHR, EMR), Existing research data (from RCTs or observational study), Other |
| Sample size | Less than 1000, between 1000 and 100,000, between 100,000 and 200,000, more than 200,000. |
| What is the study population? | Text field |
| Time range of the data for analysis | Text field |
| What were the inclusion criteria? | Text field |
| What were the exclusion criteria? | Text field |
| Was post-baseline information used to define eligibility? | Text field |
| Age range | Text field |
| **Treatment strategies and assignment** | |
| What type of RCT was emulated? | Single arm, multi-arms, Clustered |
| How many treatments were compared? | Text field |
| How were the treatment groups identified? | Based on previous RCT? Based on available data? |
| what are Treatment strategies? | Text field |
| what are the assignment procedures? | Text field |
| How were confounding factors selected? | Predefined, or statistical variable selection method |
| **Follow-up Period and Controlling Biases** | |
| How many potential time-points of the primary outcome were there? | Single, Multiple, not mentioned |
| How was immortal-time bias handled? | Text field |
| What is the maximum follow-up period? | Text field |
| Did they consider the heterogeneous effect? | Text field |
| Was selection bias due to loss to follow-up addressed explicitly? | Yes or no. |
| The statistical method used to account for potential selection bias due to loss to follow-up (if applicable). | IPTW, PGF, … |
| How was Continued Medication use assessed? | Text field |
| **Outcomes** | |
| What is the primary outcome? | Text field |
| What are the secondary outcomes? | Text field |

| | |
|---|---|
| How was the primary outcome identified? | According to the research question, randomly selected |
| **Causal contrast strategy** | |
| Which effect size measure was used to quantify the causal contrast of interest? | Mean difference, odds ratio, hazard ratio, other. |
| What statistical method was used for analyzing the primary outcome(s)? | Pooled logistic regression, Cox proportional hazards model, etc.… |
| What complementary strategies were used for results analyses? (esitmand of interest or causal contrast of interest) | ITT (Intention-to-treat) analysis, PP (Per-protocol) analysis, As-treated analysis, Other, Not Mentioned |
| **Analysis plan and statistical method** | |
| The statistical method used to account for time-varying confounders | Inverse Probability of Treatment Weighting (IPTW), Parametric G-Formula(PGF), Marginal Structural Models (MSMs), Targeted Maximum Likelihood Estimation (TMLE),… |
| What statistical methods were used for confounding adjustment? | Matching without using PS, Stratification without using PS, PS matching, Stratification on PS, Inverse probability weighting using PS (IPW / IPTW), Modeling using PS, Machine learning models, Other. |
| How was residual confounding addressed? | E-value, Negative Control, Propensity score calibration, Sensitivity analysis, Instrumental variable (IV) analysis |
| Did a new statistical model or ML propose for causal inference? | Yes, No. |
| How were ML models used to provide causal inference? | Text field |
| **Results Evaluation** | |
| Are the results identical to the results of corresponding RCT? (Is the randomization emulated well? / Are the results comparable to existing RCT?) | Yes, no, not mentioned. |
| Did they follow the TTE steps? | Text field |
| **Limitations** | |
| What were the main mentioned limitations? | Text field |
